# Protection of health care workers from exhaled air of patients operated under local, regional, spinal or epidural anaesthesia during COVID 19 pandemic

**DOI:** 10.1101/2020.11.26.20192823

**Authors:** Kuldeepsinh Pradipkumar Atodaria, Mayank Singh, Vimalkumar Prajapati, Kush Shaileshkumar Shah, Pradipkumar Raghuvirsinh Atodaria

## Abstract

The SARS-CoV-2 (COVID-19) pandemic mandates the use of N-95/FFP-2 masks for healthcare workers, especially in operation room (OR) for surgical or aerosol producing procedures. During pandemic, surgical interventions such as limb trauma, limb amputations, and limb malignancies continued to flow into the hospitals and are normally performed under local, regional or spinal anaesthesia. N-95/FFP-2 masks normally do not prevent escape of exhaled air to surrounding and to avoid the escape of exhaled unfiltered air, sealing masks by taping its edges to face possibly serves the purpose, but causes significant discomfort to patients. HEPA filters, high vacuum suction apparatus, and negative pressure operating-room may protect partially against the-risk of infection if patient’s exhaled air is infected. In order to reduce risk of transmission from patients’ exhaled air to the healthcare workers, a technique has been designed to divert the patients’ exhaled air to outside the-OR using a suction machine. This technique is easy, simple and cost-effective and trial has been performed with four-volunteers to see feasibility to breathe through N-95 mask sealed by sticking its edges to face using tape. The trial reflected reduction in SpO2, causing increased respiratory-rate, tachycardia and hypertension, in-addition an un-acclimatized volunteers had difficulty in breathing through sealed N-95 masks, which was relieved by supplying oxygen to them. Attaching suction system to remove the-exhaled air aids to comfort levels. Treating exhaled-air with sodium-hypochlorite and diverting it externally to an open-space outside the-OR added to safety for the patients, surgical team and the hospital surroundings.

## 1. Introduction

The novel coronavirus SARS-CoV-2 (COVID-19) was identified in early December in Wuhan, China (Huang et al., 2020). According to current evidence, COVID-19 virus is primarily transmitted between people through respiratory aerosol, droplets and contact routes (WHO, 2020). WHO has recommended droplet and contact precautions for Healthcare workers(HCWs) caring for COVID-19 patients, especially in settings where aerosol generating procedures and support treatment are to be performed (Lepelletier et al., 2020; Gralton et al., 2010). During the ongoing COVID-19 pandemic, performing emergency or elective surgeries carries the potential risk of infection transmission to the patients, surgeons as well as the Operating Room (OR) staff. Several surgeries are being performed under local or regional anaesthesia without closed respiratory circuits (Aliste et al., 2020).

The patients are screened pre-surgery using Reverse Transcription Polymerase Chain Reaction (RT-PCR) test which is claimed to have up to 70% sensitivity (Waller et al., 2020). 2% to 29% negative RT-PCR tests were found to be positive on repeat testing (Xiao et al., 2020). With limited availability of testing kits for COVID-19 in India at the moment, it is difficult to completely rule out COVID-19 infection in an asymptomatic patient based on one negative test. A recent Indian Council of Medical Research (ICMR) survey suggested 15-30% of patients transmitting infection, were completely asymptomatic. Asymptomatic carriers may be as infective as symptomatic patients (Chen et al., 2020). Data from various studies suggests that the virus can be transmitted through aerosol while breathing and talking. Some individual patients emit more aerosols in comparison to others during normal vocalisation (Asadi et al., 2019). Thus, operating patients under local or regional anaesthesia increases the potential risk of transmission of infection from patient to the HCWs.

In India, most patients first present to general surgeons for ailments requiring surgery, or to orthopaedic surgeons in case of trauma. These primary consultants then refer the patients to super specialists when needed. The patients are often operated upon at the premises of the primary consulting surgeon. Equipment and facilities are not necessarily same at these establishments. Hence, to protect the doctors and other HCWs in the OR from the potential infection risk from an asymptomatic infected patient, the authors came up with a customized device design. The presented device and technique are easy to replicate, delivers desirable results constantly with minimal financial implications.

## 2. Experimental

### 2.1 Innovation technique

World Health Organization guidelines recommend surgical masks for all patient care with the exception of N-95/FFP-2 masks for prolonged surgeries and aerosol generating procedures. Because of the paucity of high-quality studies in the healthcare setting, the advocacy of mask types is not entirely evidence-based (Radonovich et al., 2009). The potential benefits of respiratory protection against the possible discomfort, (Lim et al., 2006) and potential additive adverse effects on the respiratory functions of un-acclimatized patients are difficult to gauge in the absence of clear data, although there is no definite evidence of harm from prolonged usage of such masks for decades (Roberge et al., 2009). Most of plastic surgeries or cancer-reconstruction surgeries are time consuming procedures. To prevent patient’s exhaled air from leaking into the OR, the authors sealed the mask by taping it to the patient’s face.

The patients were delivered oxygen through nasal prongs, covered with a sealed N-95/FFP-2 mask to the face for preventing escape of the exhaled air. A suction cannula was also inserted underneath the mask to provide suction of exhaled air as depicted in Figure 1 (*According to the editorial instruction; Fig 1 will be provide as per the request by the readers*). The opposite end of the suction tubing is passed through two sequential suction jars containing hypochlorite solution and finally let outside the operation room. This theoretically helps to reduce the potential dispersal of virus from the exhaled air in the OR, thereby improving safety for surgical staff and at the same time provide oxygen by nasal prongs into the N-95/FFP-2 mask protect the patient from the HCWs. *The entire set up of the procedure has been provided in video file link* (*when asked for*) *for the ease of replication and adoption of the technique/devices by the other health care workers*. This technique does not compromise the patient breathing. Monitoring of the patient’s oxygen saturation, respiratory rate, pulse rate and blood pressure remains within normal limits after making the above arrangements. More elaborate studies would be needed to accurately look at the efficacy of this technique in preventing potential infection of operating room staff as well as the safety of the patient.

**Figure 1.** Positioning of the nasal prongs, suction cannula and sealing of N-95/FFP-2 mask to the face for preventing escape of the exhaled air

### 2.2 Methodology and Study design

Before implementing the device/technique for practical use in patients, the authors took a trial of the device on four volunteers after taking due consent and approval from ethics committee. Group A consisted of 2 volunteers from the medical staff who had experience of wearing N-95 masks for prolonged periods (4 hour or more). Group B consisted of regular people not accustomed to wearing N-95 masks for a long time. In all four volunteers, nasal prongs were fixed for giving oxygen, a tube for suction was also accommodated in the N95 mask before all the edges were taped to face of volunteers to prevent escape of exhaled air. The trial was consisted of three phases. In the first phase, the oxygen tubing and tube for suction were clamped. Continuous measurements of SpO2, Pulse, Respiratory rate & blood pressure were done for all four patients for a maximum of two hours or till volunteers felt symptoms of discomfort, headache, or till their SpO2 dropped below 93%. At the end of two hours, EtCO2 (End Tidal Carbon dioxide) was measured.

In the second phase of the trial, oxygen tubing were unclamped to provide oxygen through nasal prongs, but the tube for suction was not unclamped. 5 litres of humidified oxygen per minute was provided to all 4 volunteers and the same parameters were measured. The trial was given for a maximum duration of one hour or till the volunteers felt any discomfort/headache or SpO2 dropping below 93%. At the end of one hour, EtCO2 was measured.

In the third phase of trial, the suction tube was unclamped and suction machine was connected. EtCO2 was also measured in addition to the parameters mentioned in part one and two. The trial was conducted for a maximum duration of one hour or till volunteers felt discomfort/headache or SpO2 dropped below 93%. EtCO2 was measured every 15 minutes.

The total duration of our study was 4 hours, 120 minutes without O2 support, 60 minutes with O2 support but no suction, and 60 minutes with O2 support and suctioning of exhaled air.

## 3. Results and discussion

### 3.1 Observation

Two volunteers from group A and one from group B tolerated all three phases very well maintaining SpO2 > 95 %, respiratory rate and blood pressure changes within 10% of baseline, and pulse rate changes within 15% of baseline. One volunteer from group B experienced discomfort and headache at 81 minutes after SpO2 dropped below 93%, increase in pulse rate by 40%, respiratory rate by 25% and systolic BP by 20%. Oxygen tubing was unclamped and humidified oxygen at 5 litres per minute was allowed. Within 72 seconds, SpO2 became > 97% and all signs and symptoms were relieved within three minutes. EtCO2 was within normal range at the end of two hours in all four (One with oxygen). Second and third phases were tolerated very well by all four volunteers. SpO2 remained >96% in all four. Blood Pressure, RR and pulse rate were within 10% of baseline. There was no significant difference in EtCO_2_ measurements in comparison to baseline EtCO2 in all four volunteers.

### 3.2 Clinical application

After conducting the above trial and confirming the safety of the technique, we have used this technique in 27 patients so far, out of which seven were operated under local anaesthesia, fourteen under spinal anaesthesia, three under spinal epidural anaesthesia and three under brachial block. All the parameters (SpO2, respiratory rate, blood pressure, pulse rate and EtCO_2_) were measured in all patients. All patients have tolerated the N95 mask very well without discomfort except one patient who felt claustrophobia, and was kept sedated during the surgery.

All the patients were given diluted povidone iodine gargles (0.1%) before taking them to the OR (Mady et al., 2020). After this, the patients were made to wear a N-95 mask sealed with tape from the sides, provided oxygen through nasal prongs and the exhaled air was diverted using a suction tube underneath the mask. The exhaled air was passed through jars of the suction machine which contained 1% hypochlorite solution, so that the suctioned exhaled air passes through the hypochlorite solution before it is released to the external atmosphere. The machine was placed outside the OR in an open space having good ventilation.

## 4. Conclusion

The above-described technique was made to divert the exhaled air from the patient to outside the OR to avoid potential infection transmission from asymptomatic patients to health care workers (HCWs). Creating closed sealed space with N-95/FFP-2 mask with attached oxygen inlet and suction system to remove the exhaled air adds to patient comfort and prevents discomfort. The exhaled air is also chemically treated with sodium hypochlorite solution and released to an open space with good ventilation. This technique protects the patients as well as the surgical team, from infection transmission by asymptomatic carriers in OR spaces. The entire surgical team has been totally asymptomatic over a period of 10 weeks during which this technique was implemented as a standard protocol in all the OR where the authors performed surgeries.

## Data Availability

We have included all the necessary data generated during the performed surgeries. As well we followed the authors submission guidelines of the Medrxiv to put up the data fromat in manuscript.

## Notes

### Competing Interest Statement

The authors have declared no competing interest.

### Clinical Trial

CTRI/2020/11/029394

### Funding Statement

We have not received anykind of funding.

### Author Declarations

IRB/ Etics committee of Unique Hospital-multispeciality and research Institute (Ethic committee No. ECR/595/Inst/GJ/2014/RR-17 approved by Directorate General of Health Srvices, Central Drugs Standard control organization, Govt Of India)

